# Whole-exome sequencing study identifies novel rare variants and genes associated with intraocular pressure and glaucoma

**DOI:** 10.1101/2022.05.27.22275601

**Authors:** Xiaoyi Raymond Gao, Marion Chiariglione, Alexander J. Arch

## Abstract

Elevated intraocular pressure (IOP) is a major risk factor for glaucoma, the leading cause of irreversible blindness worldwide. IOP is also the only modifiable risk factor for glaucoma. Previous genome-wide association studies have established the contribution of common genetic variants to IOP. The role of rare variants for IOP was unknown. Using whole exome sequencing data from 454,756 participants in the UK Biobank (UKB), we conducted the largest exome-wide association study of IOP to date. In addition to confirming known IOP genes, we identified 40 novel rare-variant genes for IOP, such as *BOD1L1, ACAD10* and *HLA-B*, demonstrating the power of including and aggregating rare variants in gene discovery. About half of these IOP genes are also associated with glaucoma phenotypes in UKB and the FinnGen cohort. Six of these genes, i.e. *ADRB1, PTPRB, RPL26, RPL10A, EGLN2*, and *MTOR*, are drug targets that are either established for clinical treatment or in clinical trials. Furthermore, we constructed a rare-variant polygenic risk score and showed its significant association with glaucoma in independent subjects. We demonstrated the value of rare variants to enhance our understanding of the biological mechanisms regulating IOP, and uncovered potential novel therapeutic targets for glaucoma.

## Introduction

Elevated intraocular pressure (IOP) is a major risk factor for glaucoma, the leading cause of irreversible blindness worldwide. IOP is also the only modifiable risk factor for glaucoma. Current glaucoma drugs target lowering IOP. Previous studies using directly genotyped and imputed genetic data have uncovered common and some low-frequency variants for IOP^1-6^. Identifying rare variants that contribute to IOP will help uncover the biological mechanisms regulating this trait and provide improved understanding of IOP regulation and potential new therapeutic targets for managing IOP and glaucoma.

Genome-wide association studies (GWAS) have identified over 149 genetic loci associated with IOP^1-4^. These loci have established the contribution of common variants to IOP. Many of these IOP loci are also associated with glaucoma^4,5^. While these studies identified numerous loci associated with IOP, these common variants typically show small effect sizes. The role of rare variants for IOP remains to be discovered. Rare variants typically require sequencing and a large sample size to have adequate statistical power.

The UK Biobank (UKB) is a large prospective cohort of half a million adult participants with extensive genetic data linked to physical measurements, health records, family history, and lifestyle information^7^. The recent release of whole-exome sequencing (WES) data now enables the exploration of rare variants for a variety of human traits and diseases and drug targets^8,9^, including IOP and glaucoma. Rare variants can have large effect sizes and have demonstrated greater translational potential, e.g., *PCSK9*^10,11^. These WES variants are also easier to interpret because they directly map to genes.

Using WES data from 454,756 participants in UKB, we conducted an exome-wide association study (ExWAS) to identify rare variants and genes associated with IOP, evaluate their effects on glaucoma in UKB and the FinnGen cohort, and explore potential drug targets of the identified genes. We also constructed a rare-variant polygenic risk score (rvPRS) and tested its association with glaucoma. To the best of our knowledge, this study represents the largest rare-variant study of IOP to date. Our results uncovered rare variants regulating IOP, and subsequently, furthered our understanding of the biological mechanisms of IOP and potential drug targets for managing glaucoma.

## Results

We identified 14 rare variants (11 of which are novel) significantly associated with IOP, among which six were identified in white-only analysis and seven additional ones were identified in pan-ancestry analysis. Table 1 displays the single-variant association results. Our top SNP, rs74315329 (*P* = 1.22 × 10^−26^) is a well-known stop-gain variant in *MYOC*, the first gene identified for primary open-angle glaucoma (POAG)^12^. Consistently, rs28991009 in *ANGPTL7* previously identified in our array-based GWAS^2^ shows significance in this ExWAS using WES data. In white-only results, rs37278669, a nonsynonymous variant (minor allele frequency [MAF] = 0.011%), in *BOD1L1*, shows a significant association with IOP (*P* = 5.75 × 10^−9^, beta = 4.08) in UKB. *BOD1L1* is also significantly associated with the FinnGen phenotype “use of antiglaucoma preparations and miotics” (*P* = 5.10 × 10^−6^). The start-loss variant, rs753877638, in *ACAD10* is significantly associated with both IOP (*P* = 1.30 × 10^−10^, beta = 8.41, MAF = 0.003%) and glaucoma (*P* = 3.68 × 10^−4^) in UKB. In pan-ancestry analysis, rs201956837 in *HLA-B* is associated with IOP (*P* = 8.65 × 10^−9^, beta = 4.37, MAF = 0.008%). Despite rs201956837 being an intronic variant, the gene *HLA-B* is highly associated with glaucoma in FinnGen (*P* = 1.10 × 10^−9^). A Manhattan plot of the genome-wide p-values for pan-ancestry results is shown in Figure 1 (a). The genomic control lambda for white-only and pan-ancestry analyses are 1.01 and 1.02, respectively, which are well under control. The corresponding quantile-quantile plots are shown in Supplementary Figure S1.

**Table 1.**
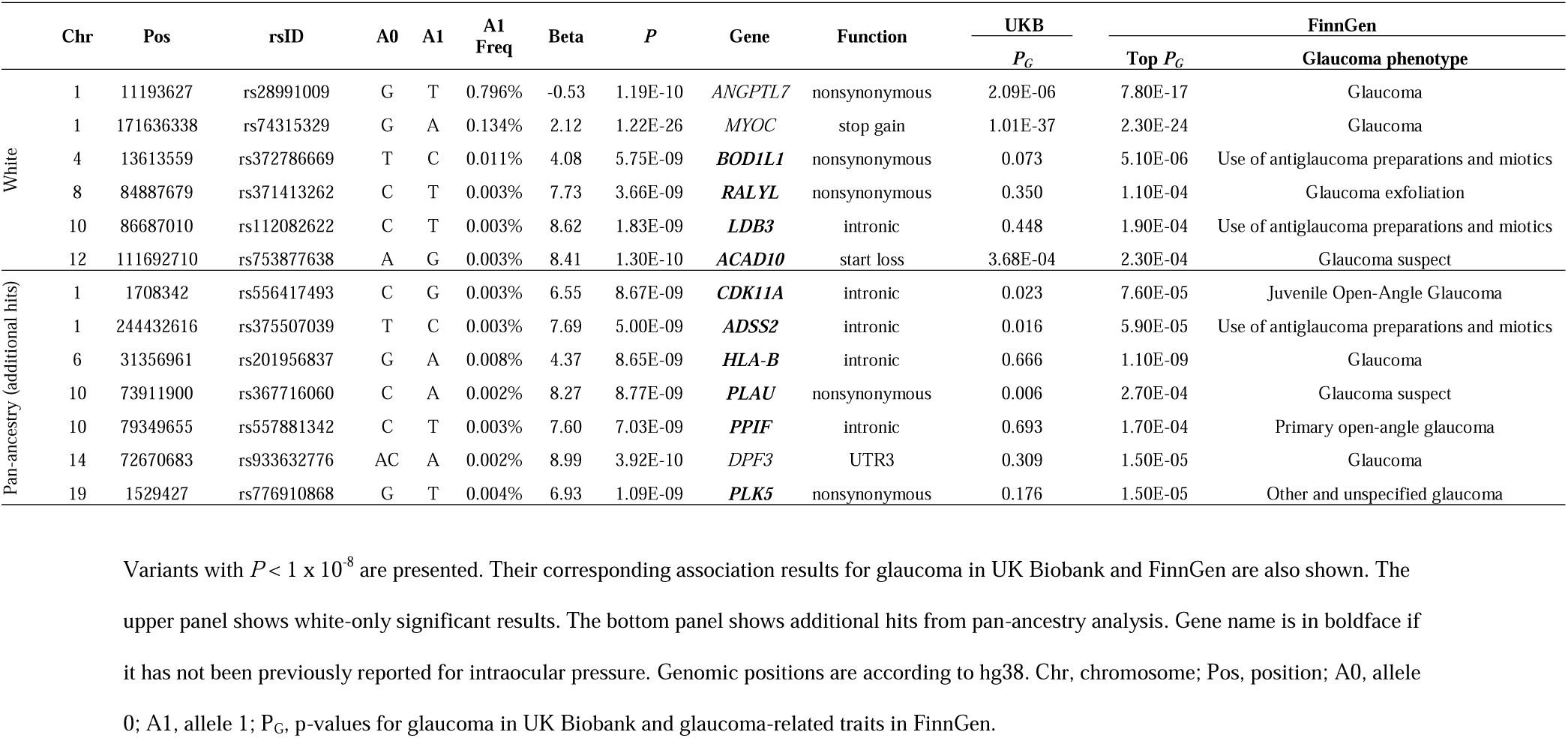
Exome-wide significant rare variants for intraocular pressure.

**Figure 1.**
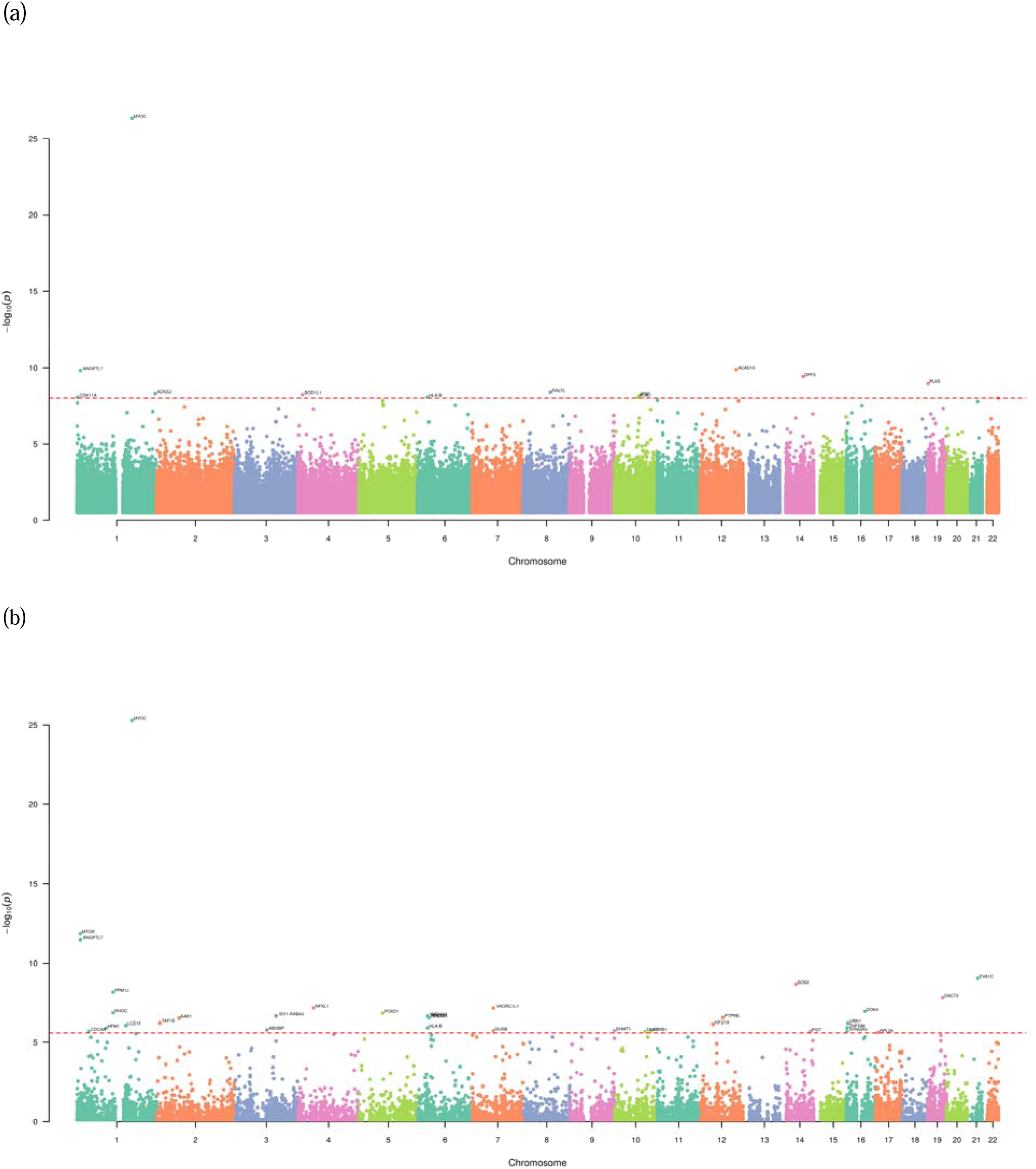
Manhattan plots displaying the -log_10_(*P*) for the association between IOP and rare variants and genes. (a) Single-variant pan-ancestry results. The dotted horizontal line represents exome-wide significance associations (*P* < 1 × 10^−8^). (b) Gene-based pan-ancestry results. The dotted horizontal line represents gene-based significance associations (*P* < 2.5 × 10^−6^). Genetic variants or genes are plotted by genomic position.

From SAIGE-GENE analysis, 35 additional genes showed significant associations with IOP and 31 of which are novel, among which 11 were identified from white-only analysis and 20 additional ones were identified from pan-ancestry analysis. Table 2 displays the gene-based association results. A Manhattan plot of the genome-wide p-values for pan-ancestry results is shown in Figure 1 (b). Rare variants in previously known IOP genes, *MTOR*^*2*^, *EVA1C*^*13*^, and *CFAP298-TCP10L*^13^, identified from common-variant investigations show significant gene-based associations with *P* = 1.08 × 10^−12^, *P* = 9.51 × 10^−10^, and *P* = 1.34 × 10^−8^, respectively. Several of these ExWAS significant IOP genes, such as *PTPRB, KIF21A, DNTT*, also show a significant association with glaucoma in UKB with *P* = 3.26 × 10^−5^, 0.009, 0.007, respectively. Many of these IOP genes are associated with glaucoma-related traits in FinnGen. For example, *PTPRB* is associated with glaucoma-related operations (*P* = 1.7 × 10^−5^), and *SYNGR3* and *ZNF598* are associated with use of antiglaucoma preparations and miotics (*P* = 4.20 × 10^−5^). *UBN1* is associated with juvenile open-angle glaucoma (*P* = 1.8 × 10^−5^). *RPL10A* and *TEAD3* are associated with exfoliation glaucoma (*P* = 9.9 × 10^−6^). *TAF1B* is associated with primary angle-closure glaucoma (*P* = 1.90 × 10^−5^). *ISY1-RAB43* is associated with normotensive glaucoma. *DOK4* is associated with glaucoma suspect (*P* = 3.20 × 10^−5^). *RHOC, PPM1J* and *LCE1F* are associated with glaucoma (*P* < 5 × 10^−5^). Notably, *ADRB1, PTPRB, RPL26, RPL10A, EGLN2*, and *MTOR* also have existing drug targets.

**Table 2.**
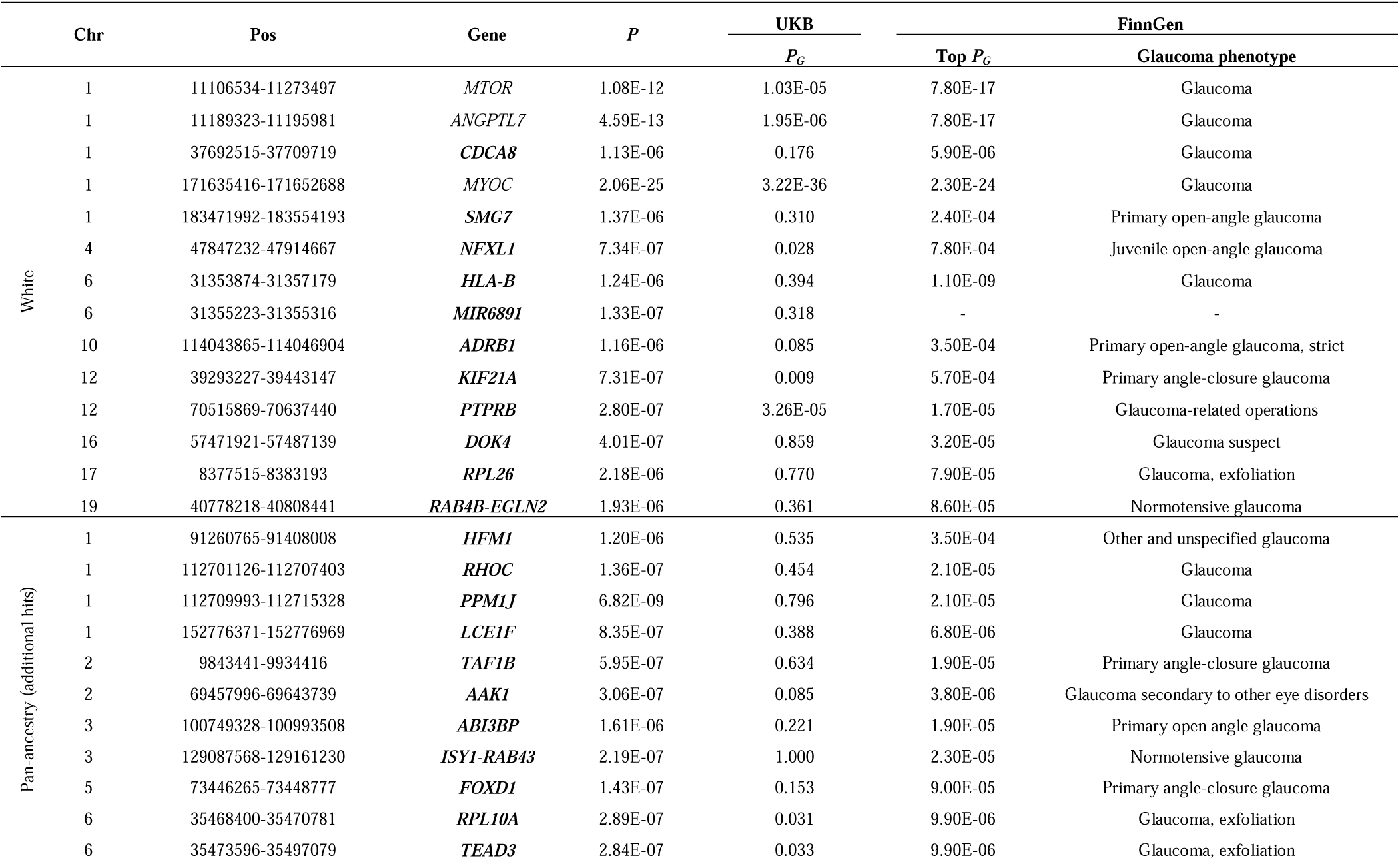
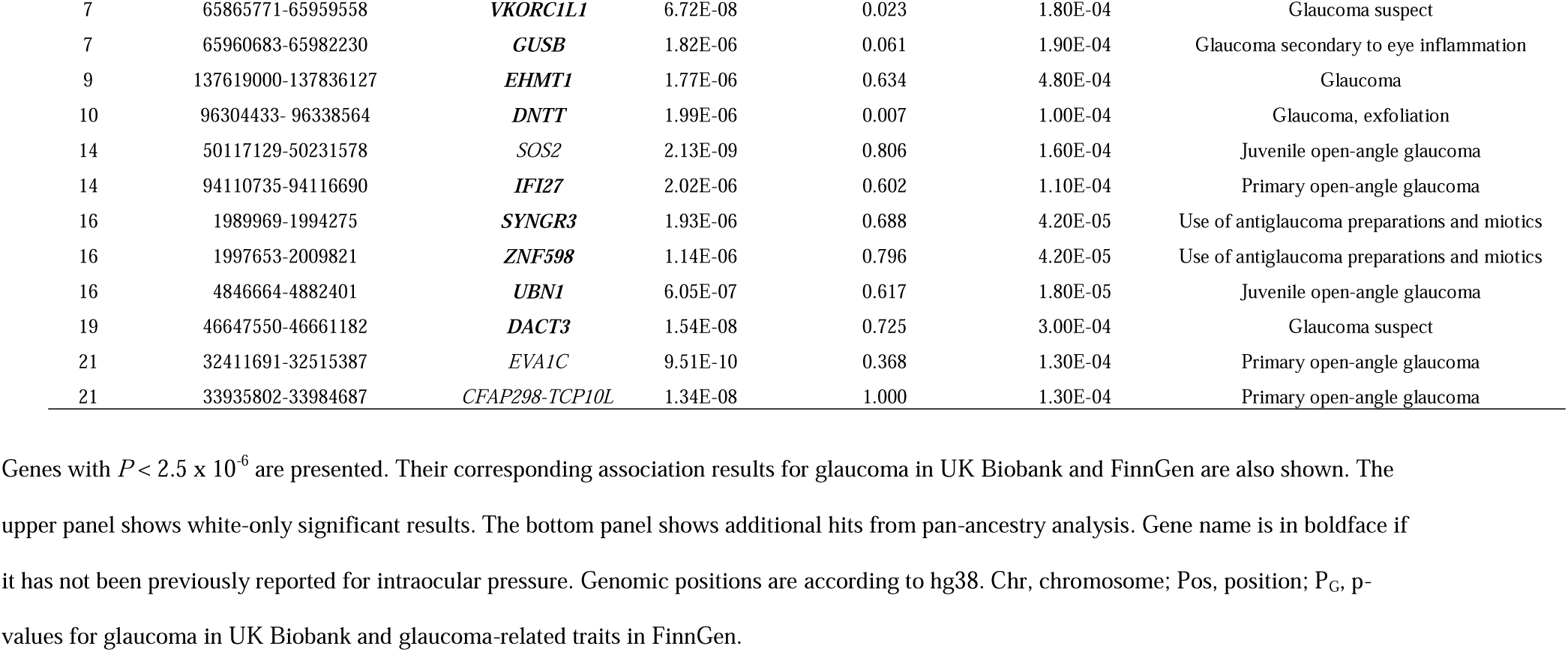
Genome-wide significant results for intraocular pressure from gene-based analysis.

Table 3 displays the current known and proposed drug targets for these IOP rare-variant genes, such as *ADRB1* (adrenoceptor beta 1), which is a known drug target for topical beta-adrenergic receptor antagonists, or beta-blockers, known to lower IOP. To the best of our knowledge, our results provided the first reported evidence for the association between IOP and *ADRB1. ADRB1* is expressed in human trabecular meshwork and ciliary body^14^, as well as cardiac tissue (Supplementary Figure S2). Glaucoma drugs targeting *ADRB1* include topical beta-blockers, such as timolol, betaxolol, carteolol, levobunolol, levobetaxolol, and metipranolol. Two older, outdated glaucoma medications include the adrenergic agonists, dipivefrin and epinephrine. In addition, several of these drugs are also used in treating hypertension and cardiovascular disease. *PTPRB* is highly expressed in the vein and artery endothelium cells (Supplementary Figure S2). It is a proposed drug target for retinal vein occlusion, diabetic retinopathy and diabetic macular edema. Razuprotafib is a small molecule targeting *PTPRB* that acts as a negative regulator of Tie2 in diseased vascular endothelium by receptor-type tyrosine-protein phosphatase beta inhibition. *EGLN2*, a neighboring gene from the readthrough gene *RAB4B-EGLN2*, has drug trials for roxadustat, daprodustat, and vadadustat, which inhibit a hypoxia-inducible factor prolyl hydroxylase. These drugs target anemia and chronic kidney disease. *MTOR* is targeted by perhexiline, a drug used for cardiovascular disease that inhibits the serine/threonine-protein kinase mTOR. The *MTOR* gene is highly expressed in microvessel endothelium cells throughout the eye (Supplementary Figure S2). *RPL26* and *RPL10A* have three experimental drugs, i.e., ataluren, ELX-02, and MT-3724, two of which (ataluren and ELX-02) work as 80S ribosome modulators while MT-3724 functions as an 80S inhibitor. These drugs are in development for various diseases, such as cystic fibrosis, muscular dystrophy, hemophilia, epilepsy, kidney disease, and leukemia.

**Table 3.**
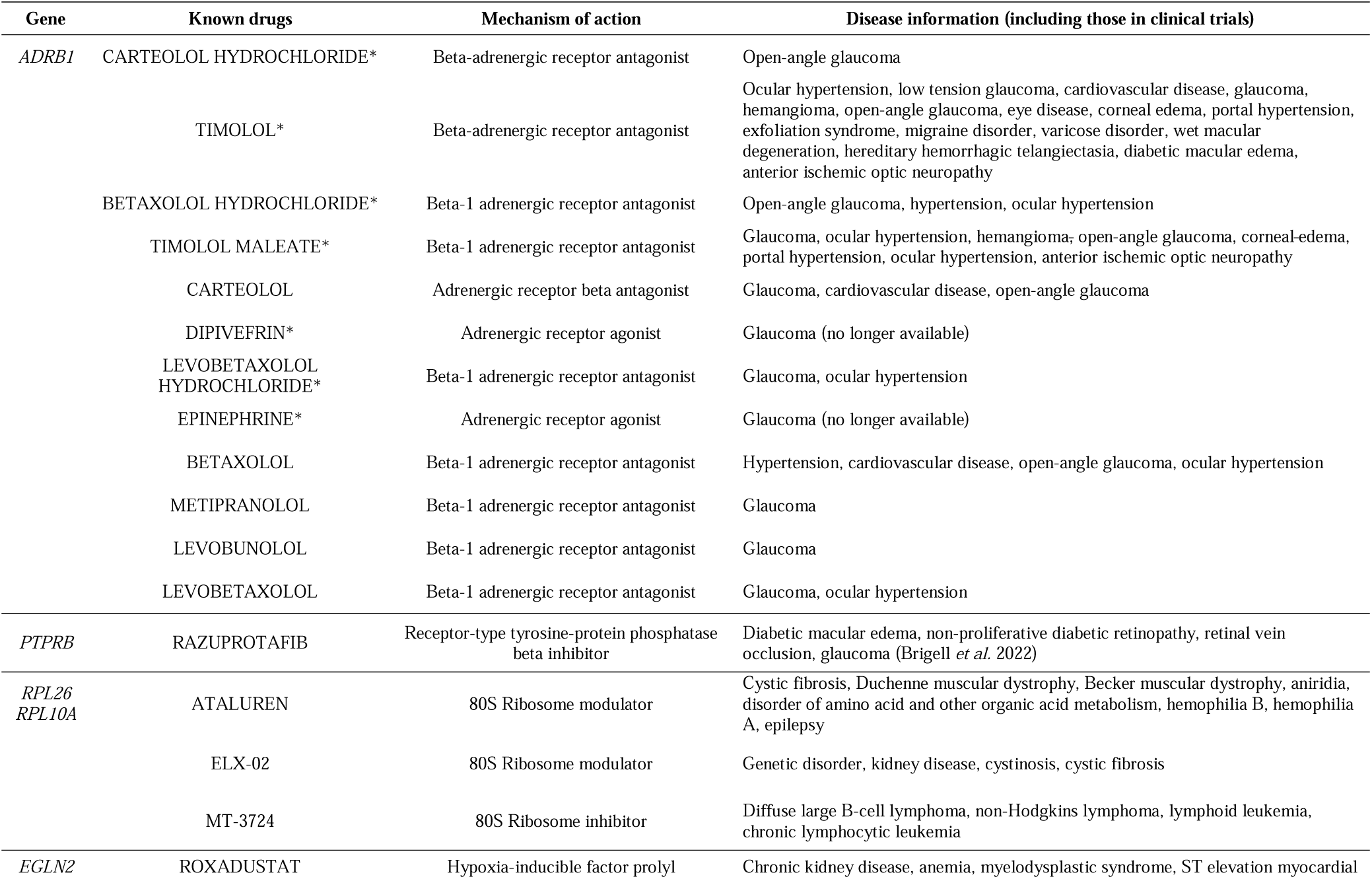
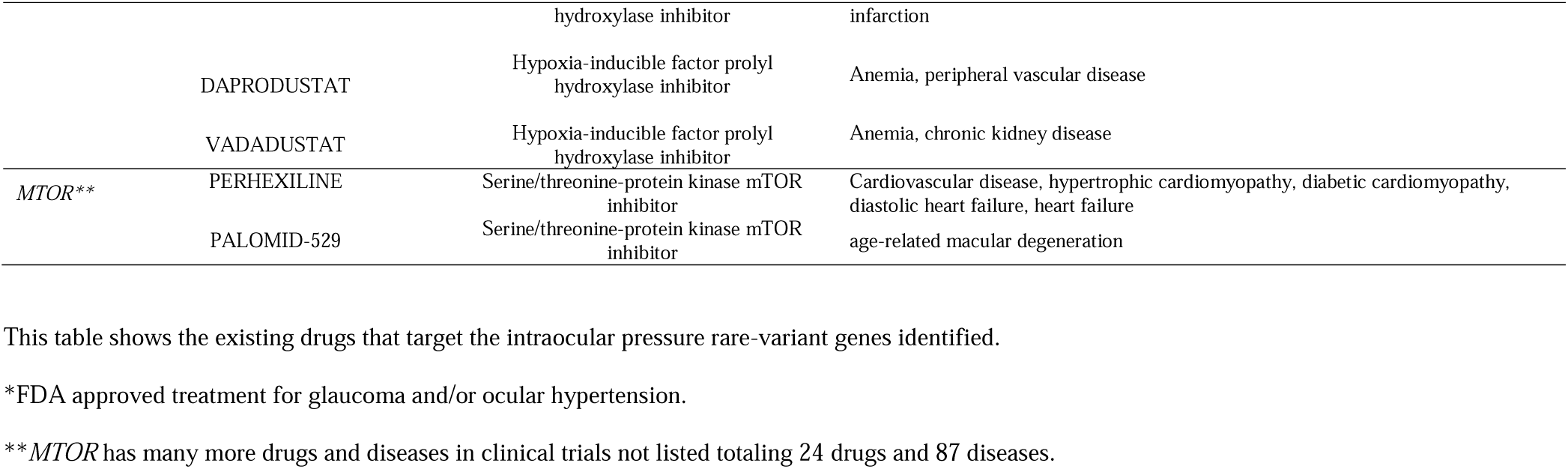
Known drug targets for the identified intraocular pressure rare-variant genes.

We further constructed a rare-variant polygenic risk score (rvPRS) using the IOP rare variants with *P* < 5 × 10^−8^ from pan-ancestry analysis (Supplementary Table S1), excluding *MYOC* due to its known large effect on glaucoma, and tested its association with glaucoma in independent UKB European subjects, who did not participate in the IOP measurements. This rvPRS is significantly associated with glaucoma with *P* = 1.02 × 10^−6^, indicating the relevance of these IOP rare variants in glaucoma. When we used the rare variants identified from white-only subjects, the rvPRS yielded a much-mitigated association with glaucoma with *P* = 6.42 × 10^−4^.

## Discussion

In this study, we conducted the largest ExWAS of IOP to date using data from UKB. By employing single-variant and gene-based analyses, two complementary frameworks, we have expanded our knowledge of the genetic architecture of IOP, especially of the role of rare variants, beyond previous studies involving microarray data, which mainly covered common variants. In additional to confirming known IOP genes, we identified 40 novel genes for IOP, demonstrating the power of including and aggregating rare variants in gene discovery. Many of these IOP genes are also associated with glaucoma phenotypes in UKB and FinnGen. Six of these genes are drug targets that are either established for clinical treatment or in clinical trials. Furthermore, we constructed a rvPRS and showed its significant association with glaucoma in independent subjects.

We also showed that including subjects of all ancestries in a pan-ancestry analysis further improved the statistical power to identify rare variants. It was evident that pan-ancestry analyses identified additional rare variants and genes beyond white-only analyses in both single-variant and gene-based analyses.

Testing these IOP variants and genes for their effects in glaucoma-related traits in both UKB and FinnGen and querying for drug targets further increased their translational relevance. Furthermore, the IOP rvPRS constructed using the rare variants identified from pan-ancestry analysis showed an even stronger association signal with glaucoma in independent white subjects than using white-only rare variants.

A concern with multi-ancestry datasets is false-positive signals. Numerous previous GWAS used European subjects only. In some studies, it was further reduced to unrelated European subjects. One way to analyze multi-ethnic GWAS datasets is using meta-analysis^1,15^, which is typically used for dealing with common variants. However, rare variants may not have enough carriers in individual ancestral groups, resulting in too few carriers to be analyzed. A pooled approach is an attractive alternative for combining ancestrally diverse populations^16^, especially for rare variants. Recent advances in statistical genetics tools also made this possible. For example, REGENIE^17^, a machine learning approach, can avoid the parameter inflation in ultra-rare-variant situations while controlling for both population stratification and sample relatedness. SAIGE^18^ uses mixed-effects models to adjust for both population stratification and genetic relationship matrix. Using these state-of-the-art methods, we showed the effectiveness of including non-European subjects in pan-ancestry analyses to further increase the study power. With advanced statistical genetics tools that can adjust for both genetic relatedness, principal components (PCs) of genetic ancestry and ancestral clusters, it is feasible to carry out pan-ancestry analyses in a pooled/combined approach. Further research is still necessary to maximize the power of pan-ancestry analysis^16^.

Genetics provides vital information to identify drug targets. The generation of this WES data is sponsored by eight pharmaceutical companies, including Regeneron and AstraZeneca^9^, which clearly shows the value of this dataset to that industry. Drug candidates that have genetics support are twice as likely to be successful than those without genetics support^19^. Six genes, i.e., *ADRB1, PTPRB, RPL26, RPL10A, EGLN2*, and *MTOR*, out of our gene-based analyses have existing therapeutic molecular targets. The most notable one, *ADRB1*, is the target of cardiovascular and glaucoma drugs, which include the broad class of glaucoma drugs targeting the beta-adrenergic receptor antagonists. For example, Timolol was a first-line drug for lowering IOP by blocking the beta-adrenergic receptors in the ciliary body^20^ to decrease aqueous humor flow^21^. More recently, timolol has been shown to have an effect on outflow facility^22^, which also impacts IOP. The other five genes are targets in many clinical trials involving razuprotafib, ataluren, ELX-02, MT-3724, roxadustat, daprodustat, vadadustat, and perhexiline, which provide candidates for drug repurposing for possible glaucoma treatment. For example, razuprotafib has been shown recently as an adjunct to latanoprost for treating glaucoma patients^23^. Razuprotafib also appears to stabilize blood vessels^23^. Roxadustat has proposed pathways affecting blood cell production^24^. Taken together, many of these drugs appear to be involved with cardiovascular disease and blood flow. Additionally, phenome-wide associations of the identified genes showed numerous significant associations with vascular-related phenotypes (Supplementary Table S2). This relationship indicates that it may be possible to repurpose certain drugs that work on cardiovascular disease for glaucoma.

This study is not without limitations. Rare variants have their intrinsic challenges. The rarity of these variants makes their replication far more difficult than common variants. Nevertheless, since IOP is an endophenotype of glaucoma and ∼70% glaucoma GWAS hits are also associated with IOP^15^, it is reasonable to test these IOP hits for their glaucoma effects^4^ although it should not be assumed that all IOP hits are associated with glaucoma. Furthermore, IOP hits can also provide translation implications for glaucoma management since lowering IOP is currently the sole proven solution for glaucoma treatment. Hence, we checked the significance of these IOP variants and genes on glaucoma-related traits in both UKB and FinnGen cohorts. In UKB, a combination of self-reported glaucoma and ICD-10/9 codes for glaucoma phenotypes is not homogeneous for specialized glaucoma subtypes, but previous studies have demonstrated the effect of using it for studying primary open-angle glaucoma genetics^4,25^. Despite being the largest WES data currently available, diversity is still low, and European subjects comprise about 94% of the UKB cohort. Other larger ancestral groups are no doubt invaluable and can provide further information for discovery and validation. About half of the IOP rare-variant genes identified were found to be associated with glaucoma-related traits in either UKB or FinnGen. Further studies are required to confirm the remaining ones for their impacts on glaucoma. Furthermore, the best approach for analyzing datasets of ancestrally diverse populations remains an ongoing research topic^16^, especially for rare variants.

In conclusion, we carried out the largest ExWAS of IOP to date. In addition to showing the efficacy of single-variant and white-only analyses, our study clearly supports using gene-based aggregation and pan-ancestry analyses to further increase the study power. We demonstrated the value of rare variants to enhance our understanding of the biological mechanisms regulating this trait, and uncovered potential novel therapeutic targets for glaucoma.

## Materials and Methods

### UKB Resource

UKB is an ongoing large prospective cohort study. Details regarding this cohort have been described elsewhere ^26,27^. Briefly, the UKB recruited over 500,000 adult participants (40 to 70 years of age at enrollment) living in the United Kingdom who were registered with the National Health Service at the study baseline (from 2006 to 2010). Medical information (self-report and electronic health records), family history, lifestyle information, as well as DNA samples, were collected. Ophthalmological data were also collected for a subset of study participants (∼118,000). Most participants (∼94%) reported their ethnic background as white and the rest originated outside of Europe^7^. Our access to the resource was approved by UKB and we obtained access to fully de-identified data.

### FinnGen Resource

The Finngen study is a large biobank study focused on the population of Finland^28^. Over 200,000 participants have been enrolled, genotyped and phenotyped. 500,000 participants are projected to be enrolled by the end of 2023. The study aims to show the power of nationwide biobanks, electronic health records and an isolated population in identifying rare variants associated with different diseases. Data was collected from different Finnish biobanks and digital health care data on Finland citizens starting in 2017. The recruited population has an age average of 63 years and hospital-based recruitment predominates thus far. Phenotypes were built using the International Classification of Disease Ninth and Tenth Revision (ICD-9 and ICD-10) codes. Genotyping was done with a custom Axiom FinnGen1 and legacy arrays and further imputed to 17 million markers based on whole-genome sequences of Finns. Out of 2,861 endpoint phenotypes created for this study, 15 are glaucoma related (Supplementary Table S3): neovascular glaucoma, primary angle-closure glaucoma, other and unspecified glaucoma, glaucoma, use of antiglaucoma preparations and miotics, juvenile open-angle glaucoma, normotensive glaucoma, glaucoma-related operations, primary open-angle glaucoma (strict), glaucoma (exfoliation), primary open-angle glaucoma, glaucoma secondary to other eye disorders, glaucoma secondary to eye inflammation, glaucoma secondary to eye trauma, and glaucoma suspect. The study used the SAIGE mixed models for their association analyses. The summary statistics are publicly online available (see web resources).

### UKB WES and Quality Control

WES for all UKB participants were generated at the Regeneron Genetic Center^8,9^. The sequencing, variant calling, and quality control were detailed previously^8,29^. Briefly, sequencing was done on the Illumina NovaSeq 6000 platform using 75 base pair paired-end reads. Variant calling and quality control were performed using the SPB protocol^30^. The high-quality WES data have been reported to exceed 20x coverage at 95.8% of targeted bases. We overlapped the data with participants who participated in the ophthalmological measurements and kept all samples that had missing rate < 2.5%. We kept autosomal variants with call rate > 95% and minor allele count (MAC) ≥ 1 (15.1 million). We annotated these variants using VEP^2^ and annovar^31^.

### IOP Measurements in UKB

IOP measurements were obtained using the Optical Response Analyzer (Reichert Corp., Philadelphia, PA) and have been described previously^32^. In brief, both corneal-compensated and Goldman-correlated IOP measurements were collected. We used corneal-compensated IOP for this study since it is less affected by corneal thickness^33,34^. The average of both eyes was used for downstream analysis. If only one IOP measurement was obtained, it was used as the final value. Study participants who received eye surgery within 4 weeks prior to the ocular assessment or those with possible eye infections did not receive IOP measurements. Moreover, we excluded study participants with extreme values of IOP, i.e., in the bottom and top 0.3 percentiles, and outliers, including subjects who had either eye surgery or used eye drop medications^2,35^. Overlapping with the WES data, 98,674 white subjects and 110,260 pan-ancestry (all ancestry combined) subjects remained.

### Single-variant and gene-based ExWAS analyses

We performed single-variant association analyses using a machine-learning method implemented by REGENIE^17^, accounting for population stratification and sample relatedness. We analyzed all variants with MAC ≥ 5 (REGENIE default) and included age, sex, and the first 10 PCs as covariates. Genetic variants with *P* < 1 × 10^−8^ were declared ExWAS significant^36^. In addition to using European subjects, recent ExWAS studies advocate to include subjects of all ancestries^37,38^. Hence, we performed both white only and pan-ancestry analyses (added an additional covariate for four major ancestral groups, i.e., European, South Asian, East Asian, and African, identified by the K-Means clustering algorithm).

For gene-based association tests, we used SAIGE-GENE^18^, a generalized mixed model approach that can adjust for both population stratification and genetic relationship. It performs rare-variant collapsing/aggregation tests, such as SKAT-O^39^, burden^40^ and SKAT^41,42^. We used predicted loss of function (pLOF) variants as the variants for gene sets. We defined pLOF variants as: stop gained, stop lost, start lost, splice donor, splice acceptor and frameshift based on the VEP^43^ annotation and gnomAD pLOF variants^44^. We included age, sex, and the first 10 PCs as covariates. Genes with *P* < 2.5 × 10^−6^ were declared significant. We performed both white-only and pan-ancestry analyses (further added dummy variables for the major ancestral groups to the covariates).

### Glaucoma lookup in UKB and FinnGen

Since lowering IOP is currently the only glaucoma treatment, we performed a lookup in glaucoma traits in UKB and FinnGen resources for all ExWAS significant IOP rare variants and genes. In the UKB subjects, glaucoma cases were identified if they self-reported glaucoma (UKB data fields 6148, 20002) or had an ICD-10 or ICD-9 diagnosis code for glaucoma (UKB data fields 131186, 131188, 41202, 41204, 41076, 41078, 41270), excluding glaucoma secondary to eye trauma, secondary to eye inflammation, secondary to other eye disorders, secondary to drugs, and other glaucoma. The selection of glaucoma based on self-reports and ICD-10 codes has been shown to be effective in previous studies^4,25^. Furthermore, the proportion of non-POAG cases in UKB was expected to be small^45^. Controls were identified as those who did not have glaucoma or self-reported eye problems. Overlapping with WES data, 14,378 white cases and 409,571 white controls, 15,606 pan-ancestry cases and 437,417 pan-ancestry controls remained. We further checked our top IOP genes in the FinnGen GWAS summary statistics^28^ by querying each of them in their online results (see web resources). For checking broader phenome-wide associations, we used PheWeb^46^.

### Drug Targets Prioritization

To prioritize drug targets for the identified rare-variant genes, we used the Open Targets online resource. For the identified genes, we queried the Open Targets for known drugs, their mechanisms of action (source ChEMBL), and disease information. The druggable genes provide key information on the relevance of these genes on IOP and glaucoma management and potential drugs for repurposing.

### rvPRS

From the pan-ancestry single-variant association results, we selected rare variants with *P* < 5 × 10^−8^ excluding *MYOC*, the well-known rare-variant gene for glaucoma. Details of these variants are shown in the Supplementary Table S1. We then constructed a rvPRS using PLINK similar to our previous approach, which was calculated as the summation of the number of rare risk alleles, assuming each risk allele has the same effect^35^. We then tested the association between the rvPRS and glaucoma in independent UKB white subjects (subjects who did not participate in the IOP measurements) using logistic regression adjusting for age and sex.

## Supporting information

All supplementary information

## Data Availability

All data produced in the present work are contained in the manuscript.

## Acknowledgements

We would like to thank the study participants from the UK Biobank as well as the staff who aided in data collection and processing.

## Web Resources

The URLs for downloaded data and programs:

ANNOVAR, http://annovar.openbioinformatics.org/

ChEMBL, https://www.ebi.ac.uk/chembl/

FinnGen, https://www.finngen.fi/

Genevestigator, https://genevestigator.com/

PheWeb, https://pheweb.sph.umich.edu/

REGENIE, https://rgcgithub.github.io/regenie/

SAIGE, https://github.com/weizhouUMICH/SAIGE

UK Biobank, https://www.ukbiobank.ac.uk

VEP, https://useast.ensembl.org/info/docs/tools/vep/index.html

