## Supplementary material for "Whole-exome sequencing study identifies novel rare variants and genes associated with intraocular pressure and glaucoma": All supplementary information

**Figure Legend**

**Supplementary Figure S1. Quantile-quantile plots for single-variant results.**

Quantile-quantile plots shows adequate control of population stratification. (a) White-only single-variant association results. (b) Pan-ancestry single-variant association results.

**Supplementary Figure S2. Gene expression for *ADRB1*, *PTPRB*, and *MTOR* from Genevestigator.**

Gene expression profiles of (a) *ADRB1*, (b) *PTPRB*, and (c) *MTOR*.

**Supplementary Table S1. Rare variants with *P* < 5 x 10^-8^ from pan-ancestry single-variant analysis.**

| **Chr** | **Pos** | **rsID** | **A0** | **A1** | **A1 Freq** | **Beta** | **Gene** | **Function** |
| --- | --- | --- | --- | --- | --- | --- | --- | --- |
| 1 | 1708342 | rs556417493 | C | G | 0.003% | 6.55 | *CDK11A* | intronic |
| 1 | 1762073 | rs536679230 | G | A | 0.002% | 8.08 | *NADK* | intronic |
| 1 | 11193627 | rs28991009 | G | T | 0.725% | -0.52 | *ANGPTL7* | nonsynonymous |
| 1 | 244432616 | rs375507039 | T | C | 0.003% | 7.69 | *ADSS2* | intronic |
| 2 | 86482074 | rs767834712 | G | A | 0.005% | 5.60 | *KDM3A* | nonsynonymous |
| 4 | 13613559 | rs372786669 | T | C | 0.010% | 4.08 | *BOD1L1* | nonsynonymous |
| 5 | 73448094 | rs567443910 | G | T | 0.010% | 3.89 | *FOXD1* | nonsynonymous |
| 5 | 75147406 | rs1396914000 | C | T | 0.003% | 7.31 | *ANKRD31* | nonsynonymous |
| 5 | 75611875 | rs1367985144 | G | T | 0.002% | 7.95 | *ANKDD1B* | intronic |
| 6 | 31356961 | rs201956837 | G | A | 0.008% | 4.37 | *HLA-B* | intronic |
| 6 | 118314013 | 6:118314013 | G | T | 0.002% | 7.99 | *SLC35F1* | intronic |
| 8 | 84887679 | rs371413262 | C | T | 0.003% | 7.72 | *RALYL* | nonsynonymous |
| 10 | 73911900 | rs367716060 | C | A | 0.002% | 8.27 | *PLAU* | nonsynonymous |
| 10 | 79349655 | rs557881342 | C | T | 0.003% | 7.60 | *PPIF* | intronic |
| 11 | 824755 | rs374358848 | G | A | 0.007% | 4.72 | *PNPLA2* | nonsynonymous |
| 12 | 111692710 | rs753877638 | A | G | 0.003% | 8.42 | *ACAD10* | startloss |
| 12 | 119516782 | rs900533998 | T | C | 0.002% | 8.14 | *CCDC60* | intronic |
| 14 | 72670683 | rs933632776 | AC | A | 0.002% | 8.99 | *DPF3* | intronic |
| 16 | 48086800 | rs749904635 | C | T | 0.003% | 7.26 | *ABCC12* | nonsynonymous |
| 19 | 1529427 | rs776910868 | G | T | 0.004% | 6.93 | *PLK5* | nonsynonymous |
| 19 | 48311573 | rs919196298 | C | T | 0.002% | 7.85 | *CCDC114* | nonsynonymous |
| 21 | 32453387 | rs143068465 | G | A | 0.981% | 0.39 | *EVA1C* | nonsynonymous |
| 22 | 50277616 | rs780142749 | G | A | 0.014% | 3.36 | *PLXNB2* | nonsynonymous |

**Supplementary Figure S1. Quantile-quantile plots for single-variant results.**

(a)

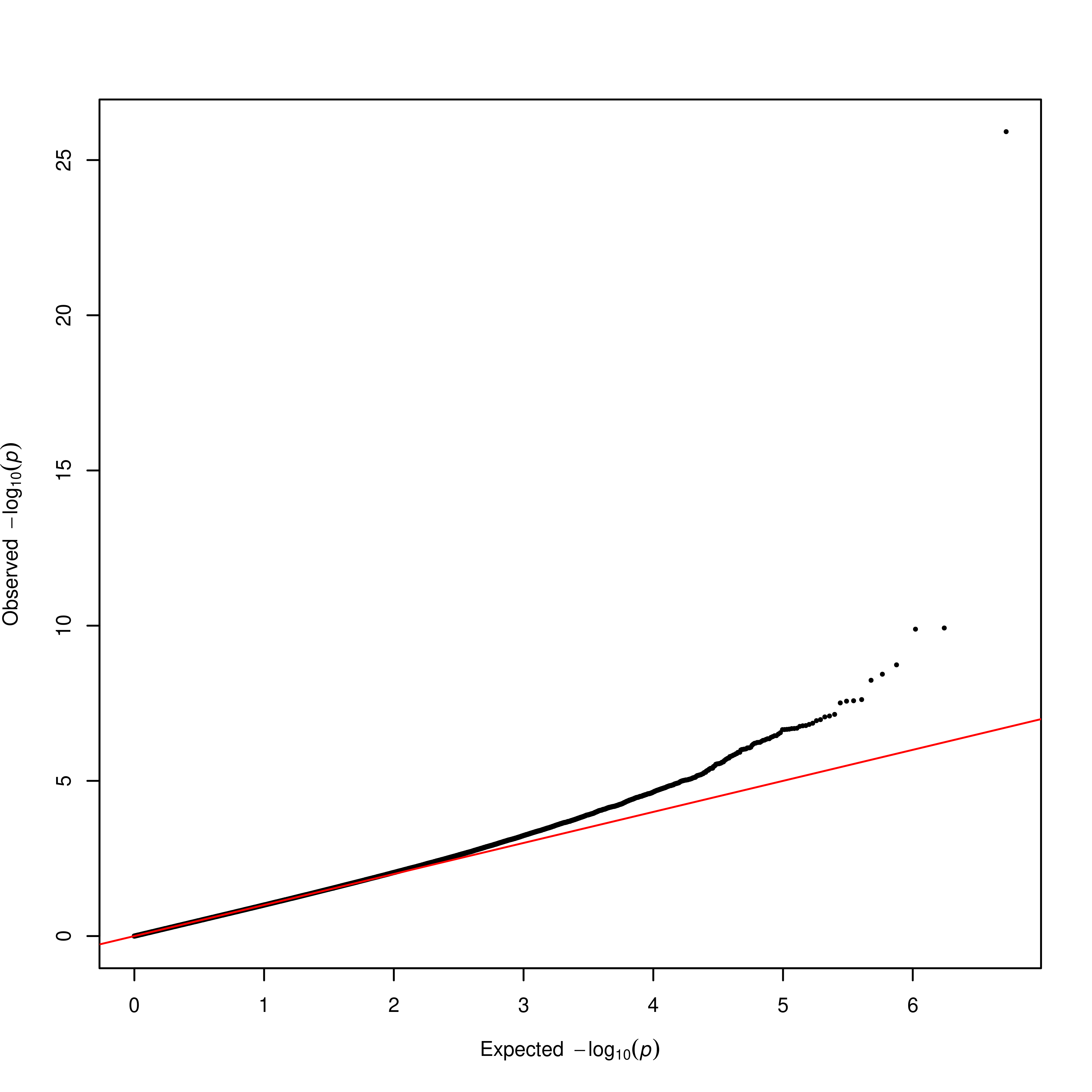

(b)

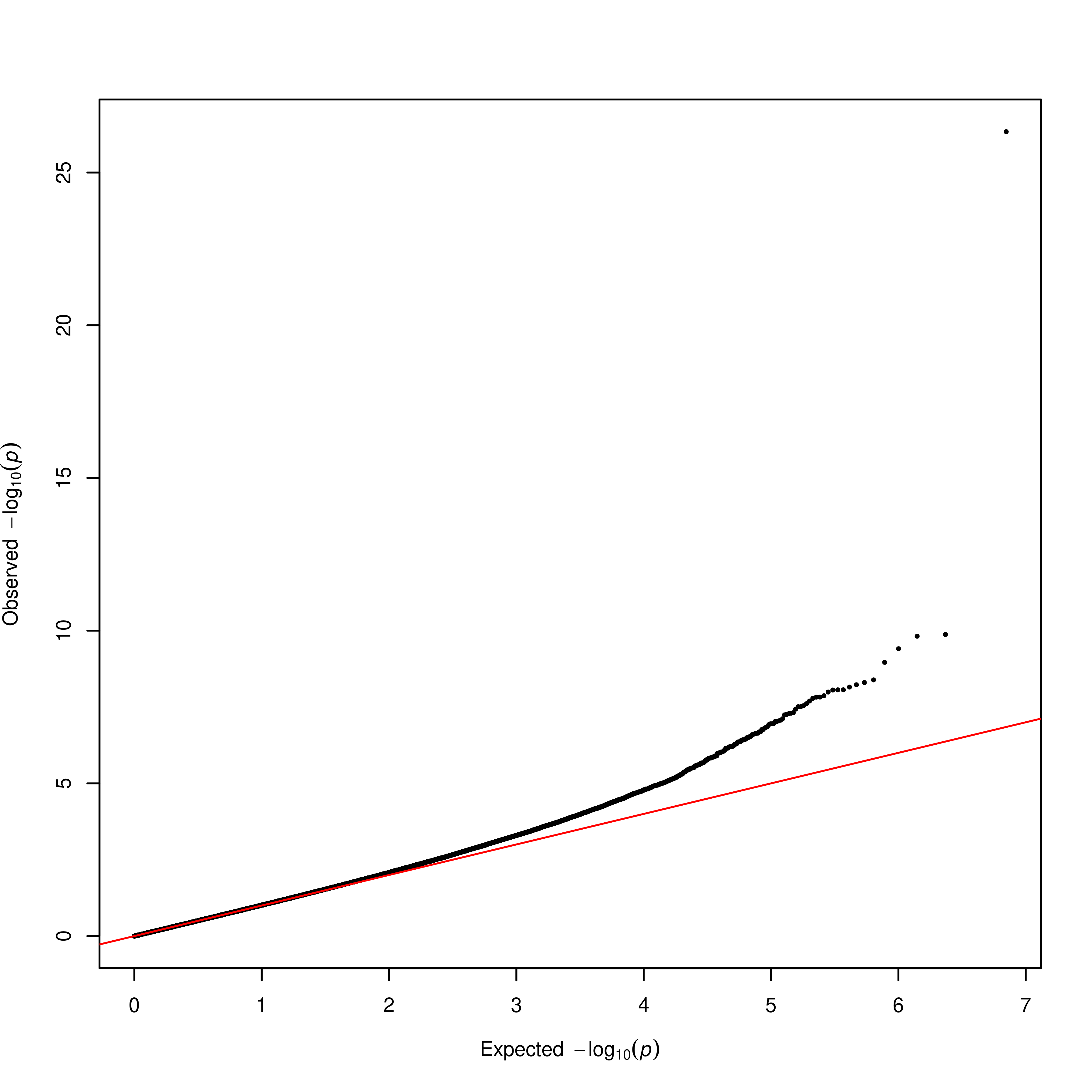

**Supplementary Figure S2. Gene expression for *ADRB1*, *PTPRB*, and *MTOR* from Genevestigator.**

(a)

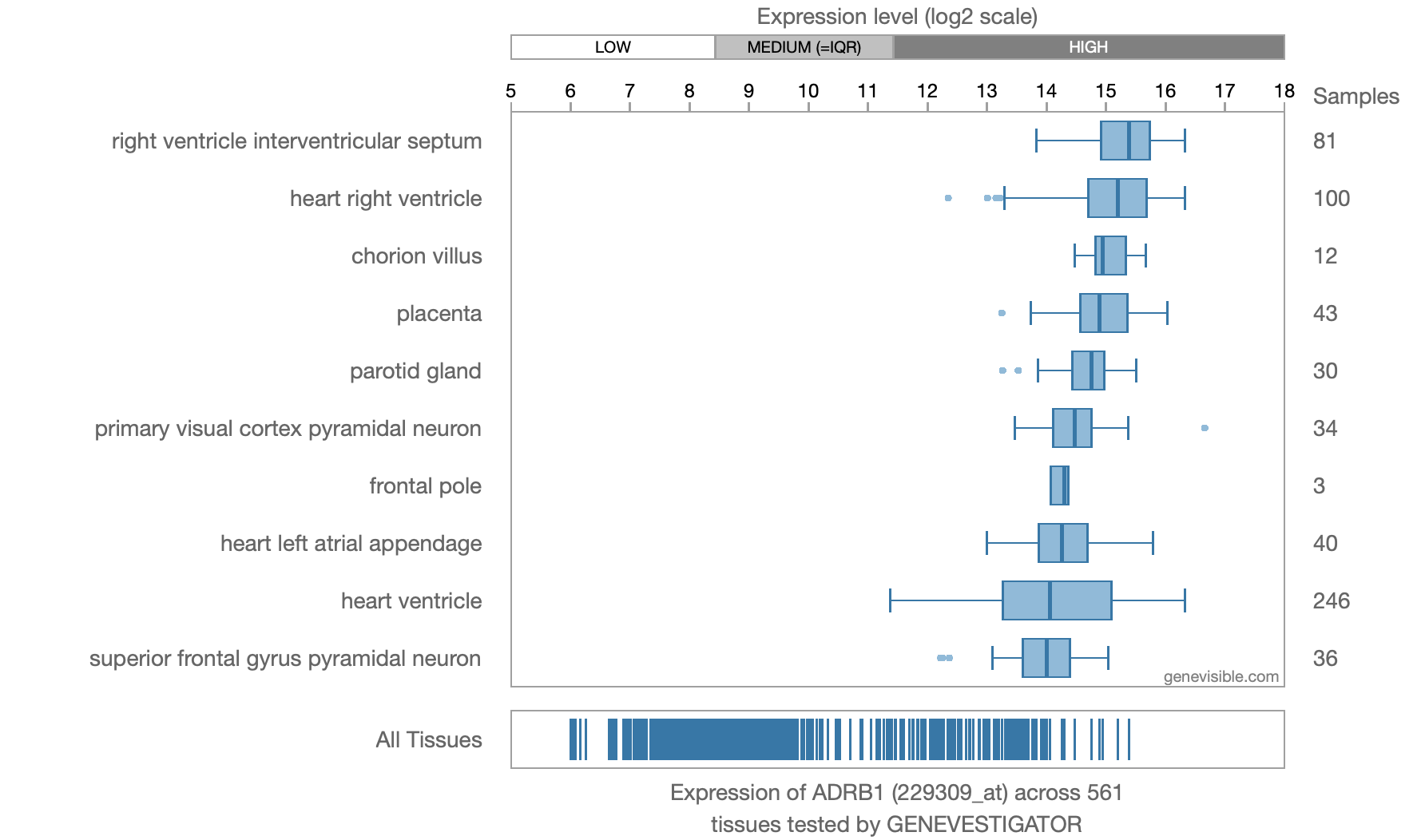

(b)

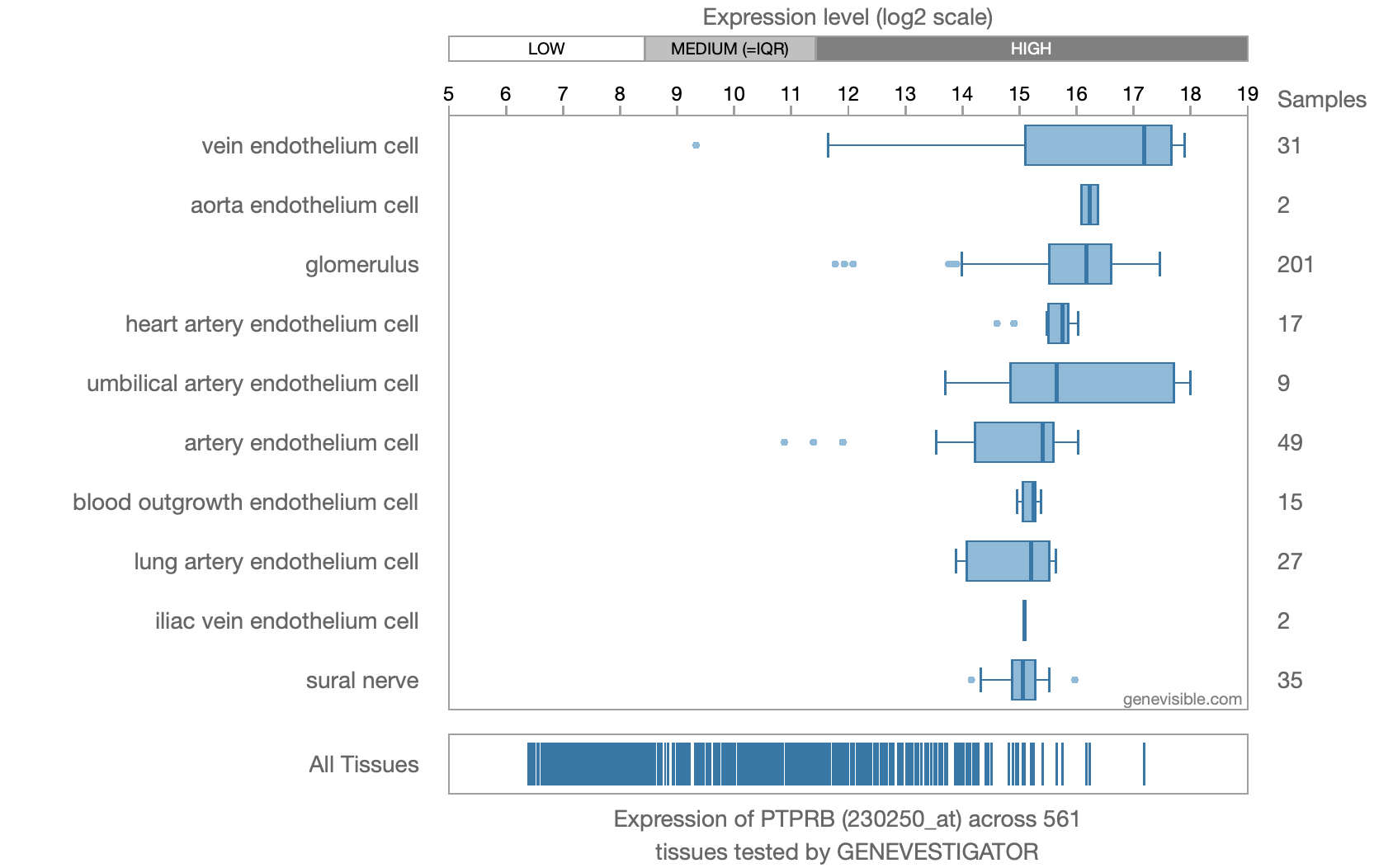

(c)

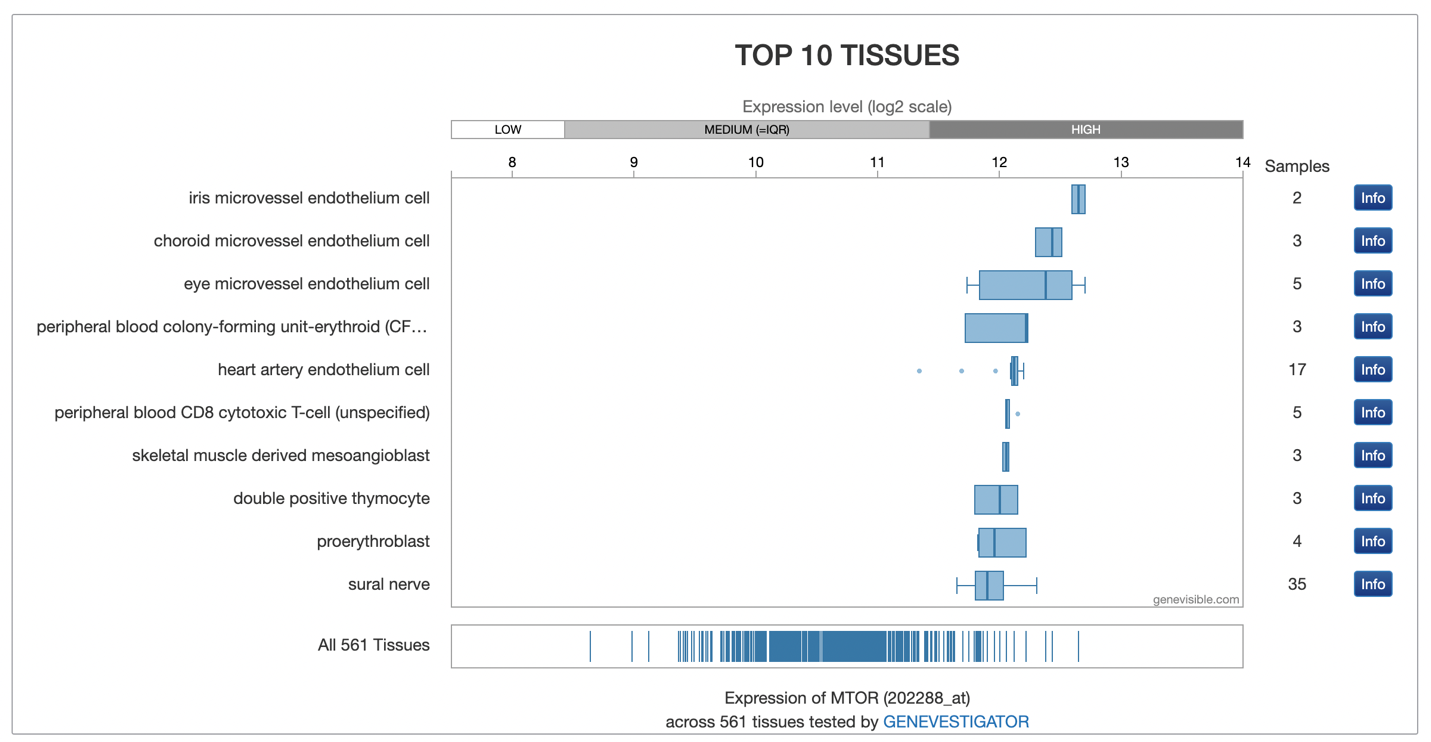

**Supplementary Table S2. PheWAS results for genes from single-variant and gene-based analyses**

| Gene | PheWeb hits  (TOPMed imputed) | *P* | PheWeb hits  (HRC imputed) | *P* |
| --- | --- | --- | --- | --- |
| *ANGPTL7* | Congenital anomaly of gallbladder | 6.50E-08 | Chronic cystitis | 8.10E-07 |
| *MYOC* | Glaucoma | 5.10E-16 | Glaucoma | 9.10E-16 |
| *BOD1L1* | Other disorders of arteries and arterioles | 1.30E-07 | Peripheral vascular disease | 5.70E-07 |
| *RALYL* | Cardiomegaly | 9.20E-07 | Chronic pulmonary heart disease | 1.50E-07 |
| *LDB3* | Essential hypertension | 6.40E-07 | Essential hypertension | 3.60E-07 |
|  | Hypertension | 8.20E-07 | Hypertension | 4.10E-07 |
| *ACAD10* | Hypertension | 2.30E-14 | Hypertension | 1.90E-14 |
| *CDK11A* | Cardiac shunt/heart septal defect | 7.30E-07 | Blindness and low vision | 2.60E-06 |
| *ADSS2* | Intracranial hemorrhage | 1.90E-07 | Acute and chronic tonsillitis | 5.00E-08 |
| *HLA-B* | Celiac disease | ≤1E-320 | Celiac disease | ≤1E-320 |
| *PLAU* | Atrial fibrillation | 7.90E-08 | Atrial fibrillation | 7.40E-08 |
|  | Chorioretinal inflammations, scars, and other disorders of choroid | 8.90E-07 | Inflammation of eyelids | 1.80E-06 |
|  | Other disorders of eyelids | 5.10E-07 |  |  |
| *PPIF* | Chorioretinal scars | 4.40E-07 | Diabetes mellitus | 1.50E-08 |
| *DPF3* | Abnormal involuntary movements | 4.50E-07 | Abnormal involuntary movements | 2.40E-07 |
| *PLK5* | Contusion | 8.10E-07 | Polyarteritis nodosa and allied conditions | 2.20E-06 |
| *CDCA8* | Respiratory insufficiency | 1.10E-07 | Aphakia and other disorders of lens | 1.00E-06 |
| *SMG7* | Diaphragmatic hernia | 1.30E-07 | Vascular disorders of kidney/hypertrophy | 1.90E-06 |
| *NFXL1* | Strabismus and other disorders of binocular eye movements | 8.50E-07 | Strabismus and other disorders of binocular eye movements | 7.10E-08 |
| *MIR6891* | - | - | - | - |
| *ADRB1* | Essential hypertension | 1.30E-15 | Essential hypertension | 3.50E-15 |
|  | Hypertension | 1.30E-15 | Hypertension | 4.50E-15 |
| *KIF21A* | Parkinson’s disease | 8.60E-07 | Tinnitus | 2.10E-07 |
|  | Disturbances of sensation of smell or taste | 6.70E-07 |  |  |
| *PTPRB* | Angina pectoris | 6.80E-07 | Type 2 diabetes | 8.60E-07 |
| *DOK4* | Coronary atherosclerosis | 3.80E-09 | Coronary atherosclerosis | 4.80E-08 |
|  |  |  | Alzheimer's disease | 8.30E-08 |
| *RPL26* | Other and unspecific disorders of the nervous system | 1.10E-06 | Contusion | 1.50E-06 |
| *RAB4B-EGLN2* | Peritoneal adhesions (postoperative) (post infection) | 8.50E-08 | Chronic tonsillitis and adenoiditis | 1.60E-06 |
| *HFM1* | Sicca syndrome | 1.10E-06 | Acquired deformities of finger | 4.10E-07 |
| *RHOC* | Hypertension | 1.30E-13 | Hypertension | 2.80E-13 |
|  | Essential hypertension | 3.10E-13 | Essential hypertension | 6.70E-13 |
|  |  |  | Optic atrophy | 9.50E-07 |
| *PPM1J* | Hypertension | 1.30E-13 | Hypertension | 2.80E-13 |
|  | Essential hypertension | 3.10E-13 | Essential hypertension | 6.70E-13 |
|  |  |  | Optic atrophy | 9.50E-07 |
| *LCE1F* | Megaloblastic anemia | 6.90E-07 | Asthma | 9.80E-07 |
| *TAF1B* | Irritable bowel syndrome | 2.10E-08 | Retinal detachment with retinal defect | 1.80E-07 |
| *AAK1* | Traumatic cataract | 7.30E-07 | Other acquired musculoskeletal deformity | 3.00E-08 |
| *ABI3BP* | Diffuse diseases of connective tissue | 2.90E-07 | Schizophrenia and other psychotic disorders | 5.80E-07 |
| *ISY1-RAB43* | Blindness and low vision | 8.70E-07 | Chronic periodontitis | 2.30E-07 |
|  | Other and unspecific disorders of the nervous system | 6.90E-07 |  |  |
| *FOXD1* | Benign neoplasm of eye, uveal | 5.50E-07 | Other chronic nonalcoholic liver disease | 2.10E-08 |
| *RPL10A* | Atherosclerosis of aorta | 3.40E-07 | Celiac disease | 1.00E-12 |
| *TEAD3* | Atherosclerosis of aorta | 3.40E-07 | Celiac disease | 1.00E-12 |
| *VKORC1L1* | Other inflammatory spondylopathies | 1.60E-07 | Cancer of eye | 1.90E-06 |
| *GUSB* | Other inflammatory spondylopathies | 1.60E-07 | Cancer of eye | 1.90E-06 |
| *EHMT1* | Subjective visual disturbances | 4.50E-07 | Aneurysm of artery of lower extremity | 1.00E-06 |
|  |  |  | Hypertrophic obstructive cardiomyopathy | 1.40E-06 |
| *DNTT* | Anterior horn disease | 3.40E-07 | Appendiceal conditions | 5.70E-07 |
| *SOS2* | Hypertension | 1.70E-09 | Hypertension | 9.50E-09 |
|  | Essential hypertension | 2.30E-09 | Essential hypertension | 1.50E-08 |
| *IFI27* | Visual field defects | 1.30E-07 | Emphysema | 1.10E-07 |
| *SYNGR3* | Atrial fibrillation and flutter | 4.80E-09 | Atrial fibrillation and flutter | 5.40E-09 |
|  | Aneurysm of other specified artery | 9.20E-08 | Cardiac dysrhythmias | 3.00E-07 |
|  | Cardiac dysrhythmias | 1.90E-07 |  |  |
| *ZNF598* | Atrial fibrillation and flutter | 4.80E-09 | Atrial fibrillation and flutter | 5.40E-09 |
|  | Aneurysm of other specified artery | 9.20E-08 | Cardiac dysrhythmias | 3.00E-07 |
|  | Cardiac dysrhythmias | 1.90E-07 |  |  |
| *UBN1* | Phobia | 3.60E-08 | Fracture of humerus | 4.90E-07 |
| *DACT3* | Opiates and related narcotics causing adverse effects in therapeutic use | 1.20E-07 | Vascular disorders of skin | 5.60E-06 |
|  |  |  | Other conditions of brain | 5.60E-06 |
| *EVA1C* | Ptosis of eyelid | 8.30E-07 | Arthropathy NOS | 3.60E-07 |
|  | Cardiac dysrhythmias | 1.10E-06 |  |  |
| *CFAP298-TCP10L* | Ptosis of eyelid | 8.30E-07 | Epiphora | 7.40E-07 |
|  | Von willebrand disease | 7.80E-07 |  |  |

The results from phenome-wide association study (PheWAS) on the genes found in rare-variant and gene-based analyses are shown. From PheWeb top hits list, we extracted eye related, cardiovascular and nervous system related phenotypes if available, otherwise the top hits were extracted.

**Supplementary Table S3. FinnGen glaucoma phenotypes and their extraction criteria.**

| **FinnGen glaucoma phenotypes** | **Endpoint definitions** |
| --- | --- |
| Primary angle-closure glaucoma | H40.2 (ICD-10), 3652 (ICD-9), 37500\|37511 (ICD-8) |
| Other and unspecified glaucoma | H40.8\|H40.9 (ICD-10), 3656X\|3656C\|365[8-9] (ICD-9), 3752\|3759 (ICD10-8) |
| Glaucoma | H40\|H42 (ICD-10) |
| Use of antiglaucoma preparations and miotics | Medicine purchases: ATC S01EA, Min. number of events: 3 |
| Juvenile Open Angle Glaucoma | Pre-conditions 4<=EVENT_AGE<=40 |
| Normotensive glaucoma | H40.11 (ICD-10), 3651C (ICD-9) |
| Glaucoma-related operations | Operations: NOMESCO codes ^CHD15\|^CHD50\|^CHD65\|^CFW99 |
| Primary open-angle glaucoma, strict |  |
| Glaucoma, exfoliation | H40.10 (ICD-10), 3651B (ICD-9), 37512 (ICD-8) |
| Primary open-angle glaucoma | H40.1 (ICD-10), 3651 (ICD-9), 3751[0-3] (ICD-8) |
| Glaucoma secondary to other eye disorders | H40.5 (ICD-10), 3655\|3656A (ICD-9) |
| Glaucoma secondary to eye inflammation | H40.4 (ICD-10), 3656F\|3656B (ICD-9), 36401 (ICD-8) |
| Glaucoma secondary to eye trauma | H40.3 (ICD-10), 3656E (ICD-9) |
| Glaucoma suspect | H40.0 (ICD-10), 3650 (ICD-9), 37597 (ICD-8) |

Source of endpoint definitions: https://risteys.finngen.fi/
